# Treatment of thoracic outlet syndrome to alleviate chronic migraine headache

**DOI:** 10.1101/2023.08.29.23294801

**Authors:** L Randall, S. Ahn, J. Weber, Y.H. Cha

**Affiliations:** Mayo Clinic Alix School of Medicine, Rochester, MN; Texas Christian University, School of Medicine, Fort Worth, TX; Dallas Fort Worth Vascular Surgery, Dallas, TX; St. Francis Medical Center, Tulsa, OK; Department of Neurology, University of Minnesota, Minneapolis, MN

**Keywords:** thoracic outlet syndrome, headache, migraine, brachial plexus, subclavian vein

## Abstract

**Background:** Prior case reports have indicated that treating thoracic outlet syndrome (TOS) may relieve intractable migraine headaches, but the reported experience has been limited. We report in a large case series how a strategy of treating concurrent TOS can help relieve intractable migraine headaches in patients with these dual diagnoses.

**Methods:** Retrospective chart review for diagnostic and interventional data on patients with migraine and TOS followed by a questionnaire to investigate specific migraine features and changes in headache burden before and after treatment of TOS.

**Results:** 50 patients (48 women, 2 men, age = 43.9+/12.7years) with dual diagnoses of chronic migraine and TOS were included (20 migraine with aura, 28 migraine without aura, two hemiplegic migraines). Based on review of available data, headaches had become chronic within one year of onset in 21 patients (42%) and included these characteristics: side-locked or greater severity ipsilateral to limb paresthesia (38/50 patients), presence of limb swelling (32/48 patients), and worsened by recumbency (32/38 patients). Interventions included physical therapy, percutaneous transluminal venoplasty, 1st rib removal, scalenectomy, pectoralis minor tenotomy, and vein patching. Thirty-two patients needed surgery. Mean patient-reported improvement of headaches on the treated side was 72+/-26.7%; 12 patients experienced complete resolution of headaches after surgical treatment of TOS (follow-up 7.2+/-5.2 months). Questionnaire responders reported significant reductions in headache days (18.3+/-8.6 to 11.1+/-10.8 days/month, p<0.0016), severity (7.8+/2.5 to 5.4+/-2.9, p<0.00079), and need for emergency care (3.6+/-4.0 to 0.71+/-1.3 visits/year, p<0.0029) after having had their TOS treated with surgery. Questionnaire responders and non-responders were not significantly different in underlying clinical features.

**Conclusion:** Chronic migraines can be important manifestations of TOS. Early transition to a chronic state, headaches worsened by recumbency, and headaches with lateralized myofascial pain are clues to a contribution by TOS pathology. The TOS contribution to migraine has been under-recognized. Addressing it can significantly improve migraine headache burden.

## INTRODUCTION

Migraine headaches lead to tremendous disability and loss of productivity, particularly when they become intractable and therapeutic options have been exhausted ^1^. While migraine is considered to be a primary headache disorder and should be diagnosed when explicit criteria as outlined in the International Classification of Headache Disorders (ICHD) are met, non-headache-related symptoms may provide clues that a structural cause may be a contributor ^2–4^. An underrecognized contributor to migraine is compression of the brachial plexus, subclavian artery, and/or subclavian vein leading to thoracic outlet syndrome (TOS) ^5–7^. In patients whose migraines are secondary to TOS, the associated neck and limb pain should be recognized as clues of a structural contributor.

Previous studies have demonstrated that migraines can be a prominent symptom of TOS but worldwide experience has been limited ^8–11^. In 1985, Raskin reported on 30 patients with TOS whose migraine headaches preceded the onset of arm symptoms and had significantly improved headache control after treatment of TOS ^8^. Given the era, there had been no imaging performed to diagnose vascular TOS but contemporary criteria for neurogenic TOS had been met. In 1999, Saxton and colleagues reported detailed anatomical imaging with MRI and MRA of the brachial plexus in a 36-year-old woman with childhood-onset migraine headaches whose premonitory symptoms of the migraine headache were bilateral hand pain and paresthesia ^10^. Her imaging showed bilateral cervical ribs and an aberrant subclavian artery compressing the vagus and recurrent laryngeal nerves, the axillary-subclavian vein, and the stellate ganglia. The patient’s migraines resolved with unilateral 1^st^ rib resection, middle and anterior scalenectomy, and extensive brachial plexus neurolysis. In 2017, Chahwala detailed a case showing retrograde flow of contrast from the subclavian veins to the internal jugular vein on arm adduction with enhanced collateralization in a patient with chronic migraine ^9^. The migraines were relieved by scalenectomy and 1^st^ rib resection. Despite the prominence of myofascial pain and lateralized motor and sensory symptoms in migraines, recognition of TOS as an underlying contributor to intractable migraines has been almost completely overlooked since Raskin’s report nearly 40 years ago.

TOS is designated according to whether the subclavian vein, subclavian artery, or brachial plexus is affected. Neurogenic TOS (nTOS) relates to symptoms caused by compression of the brachial plexus and manifests with paresthesia, numbness, pain, and weakness of the affected arm and hand ^12^. Arterial TOS (aTOS) relates to compression or injury of the subclavian artery leading to symptoms such as pain and pallor due to ischemia, including the production of emboli to the hand. It isusually associated with cervical ribs or bony abnormalities in the thorax ^13, 14^. Venous TOS (vTOS) relates to compression or thrombosis of the axillary or subclavian vein; it may present as McCleery syndrome when there is no thrombosis or as Paget Schroetter’s syndrome, i.e.“effort thrombosis,” when there is an associated deep vein thrombosis^15, 16^. Venous compression leads to swelling, warmth, and erythema. Compression of these thoracic outlet structures frequently occurs together, however, making it difficult to distinguish symptoms due to each structure ^5, 17^. For example, subclavian vein compressions can occur in as many as 65% of patients with neurogenic TOS ^11, 18^.

We took on this study to determine which clinical features should alert the clinician of when TOS is contributing to migraine headache burden. This is particularly important because many of the patients in this series had experienced severe migraine headaches for years, including starting in childhood, and only had headache relief once the TOS was treated. While most data presented is from chart review, patients with a dual diagnosis of chronic migraine and TOS were asked to provide more detailed information about treatment effects in the form of a questionnaire so that more specific features could be ascertained. The recognition of TOS contributing to migraine headaches can be critical in converting intractable headaches to manageable ones and sometimes in completely relieving them.

## METHODS

### Participant inclusion criteria

Patients experiencing chronic migraines defined by ICHD-3 criteria as ≥15 days of headaches per month in which at least 8 are migraine headaches lasting more than 4 hours a day for ≥3 months were included in the study ^2^. Patients could also meet the criteria for New Daily Persistent Headaches (NDPH) as a secondary diagnosis. The entry criteria for TOS were based on nTOS since these are the most clearly defined criteria by the Consortium for Education and Outcomes Research on Thoracic Outlet Syndrome (CORE-TOS) and the Society for Vascular Surgery (SVS) ^19^ and can be diagnosed by clinical history and exam ^19^. This includes at least 4 of 5 clinical features being met: 1] pain or paresthesia in the neck, upper back, shoulders, arm, and/or hands, 2] symptoms exacerbated by arm elevation, repetitive use, or radiation of pain/paresthesia from the supra or infraclavicular regions, 3] a mechanism for injuries such as overuse, or prior trauma to the head, neck or chest, 4] tenderness over the scalene muscles or subcoracoid areas, 5] a positive upper limb tension test or elevated arm stress test ^19^. In addition, concurrent vTOS was evaluated with CT chest with contrast, digital subtraction venography, and evaluation for the presence of hand and arm swelling ^20^ (**Figure 1**). All patients in this study met criteria for both nTOS and vTOS.

**Figure 1:**
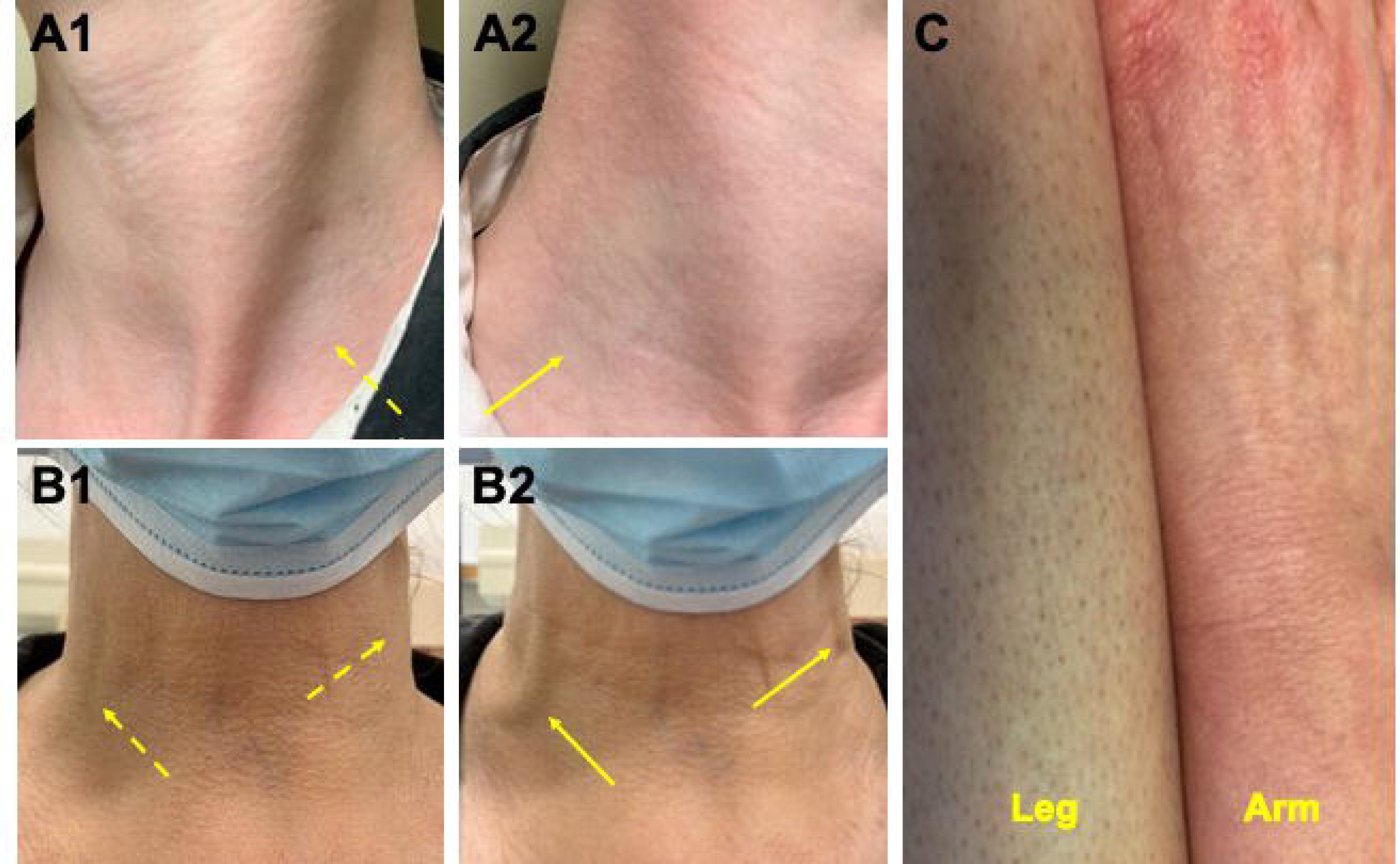
Examples of clinical signs of venous obstruction. A1] Left supraclavicular fossa, dashed arrow showing normal concavity. A2] Right supraclavicular fossa, solid arrow showing loss of definition of the lower sternocleidomastoid posterior border due to swelling in a patient with right vTOS. B1] Arms at the sides, dashed arrows showing flat external jugular veins. B2] Arms abducted to 90 degrees, solid arrows showing dilated external jugular veins in a patient with bilateral vTOS. C] Flushness and swelling of an arm in a patient with bilateral vTOS (only one arm shown), compared to the normal skin tone of the leg.

Patients underwent CT chest with contrast injected into the symptomatic arm to determine the presence of subclavian vein thrombosis, stenosis between the clavicle and 1^st^ thoracic rib, cervical ribs, or structural chest abnormalities such as pectus excavatum and axillary masses. This was followed by digital subtraction venography in patients who required quantitative assessment (**Figure 2A-C**). Arterial TOS was evaluated by chest CT ruling out cervical ribs and by arteriography when there was suspicion of arterial involvement. MRI cervical spine, MRI shoulder, and EMGs were performed to resolve diagnostic ambiguity when there was a suggestion of cervical radiculopathy, ulnar, or median neuropathy. MRI or CT of the brain was performed to rule out other secondary causes of headache.

**Figure 2:**
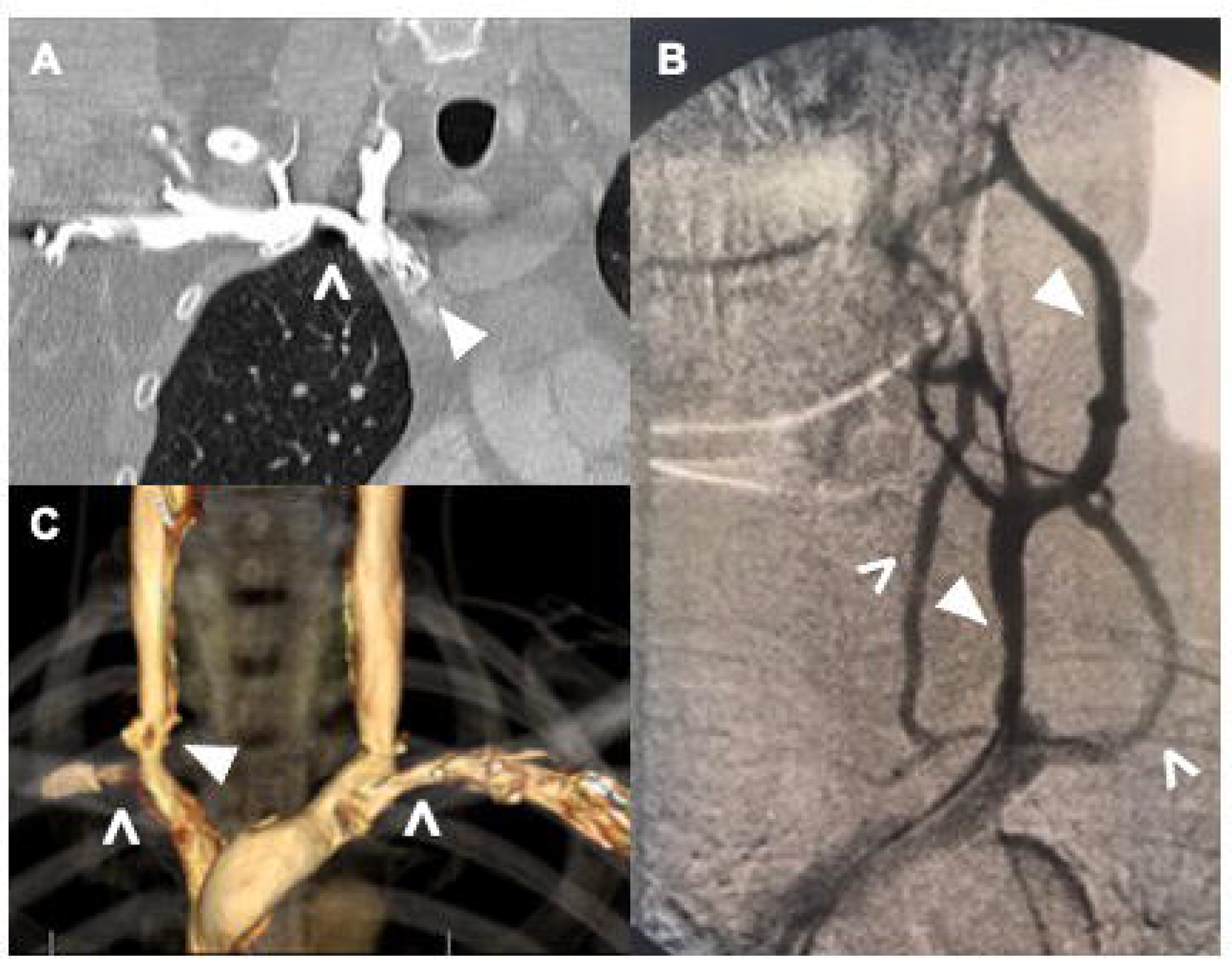
Representative examples of imaging in this cohort of chronic headache and TOS patients. Relevant abnormalities are marked by either open arrowheads or closed arrowheads. A] Right subclavian vein narrowing between the clavicle and 1^st^ rib (open arrowhead) with contrast reflux into the internal jugular vein and lack of flow into the innominate vein (closed arrowhead), B] Overdeveloped external vein (closed arrowheads) with extensive collaterals (open arrowheads) in the setting of a stenosed subclavian vein. C] 3D reconstruction of venous system showing bilateral subclavian vein narrowing (open arrowheads) as well as a narrowed right-sided internal jugular vein (closed arrowhead).

Patients in whom TOS was identified underwent an escalating series of treatments including physical therapy, percutaneous transluminal venoplasty (PTV), and surgical decompression including 1^st^ rib resection plus scalenectomy, scalenectomy alone, pectoralis minor tenotomy, or vein patching as recommended by the consulting vascular surgeon. Patients with limb swelling were treated with diuretics. Medical management with standard headache prevention protocols continued through the TOS treatment course for as long as the patient presented for follow-up. Improvement in headaches was attributed to the treatment of TOS by temporal association after specific intervention for TOS.

A chart review of all patients seen by the principal investigator over the course of 3 years for a dual diagnosis of chronic migraine and TOS was performed with data extracted as the patients were being followed. Institutional approval to send questionnaires outside of regular clinical care was obtained from the Saint Francis Health Systems Institutional Research Ethics Board with protocol number 2250-19. Informed consent was obtained from all participants with research performed in accordance with the Declaration of Helsinki. Participant surveys were conducted to assess for more detailed TOS and headache features, therapies attempted, changes in symptoms following treatment, and general well-being and quality of life. Patients were sent a SurveyMonkey® weblink with a unique study identifier that only they could access.

### Measures

Descriptive data on headache time course and diagnoses are reported on all patients for which that information could be reliably obtained. All post-treatment values were compared against pretreatment values (e.g. headache days before surgery vs. after surgery). Statistically significant changes in metrics such as the average number of headache days, severity of headaches, and number of yearly urgent care/emergency room visits before and after treatment of TOS were calculated as two-tailed paired t-tests with a significance of *p*<0.05. Data are presented for the number of participants who had sufficient information for that measure with the number of data points reported. Therefore, some data reported have different denominators.

## RESULTS

### Clinical features

A total of 50 patients from a base of approximately 600 patients referred for headaches met the criteria for both chronic migraine and TOS between March 2016 and April 2019. Clinical features of 48 women and two men (age 43.9+/-12.7 years, median 44 years) are detailed in **Table 1**.

**Table 1:** Timeline and diagnosis of chronic migraine TOS patients.

| **Headache time frame (n=50)** | *Mean +/- s.d.* | *Median (range)* |
| --- | --- | --- |
| Age at presentation | 43.9+/-12.7 years | 44 (17-78) years |
| Age at onset any migraine | 23.9+/-14.9 years | 23.5 (5-62) years |
| Age at onset chronic migraine | 34.8+/-13.4 years | 36 (5-62) years |
| Duration chronic migraine | 9.3+/-12.7 years | 5.8 (0.3-73) years |
| **Headache types (n=50)** |  |  |
| Migraine without aura total |  | 28 |
| (+) NDPH* |  | 7 |
| Migraine with aura total |  | 20 |
| (+) NDPH* |  | 1 |
| Hemiplegic migraine |  | 2 |
| Chronic migraine within 1 year of headache onset |  | 21 |
| *New Daily Persistent Headache |  |  |

Headaches were unilateral and side-locked in 17 patients, and when bilateral were worse on the side of the greater arm symptoms in an additional 21 patients (**Table 2**). Thirty-two patients endorsed swelling in their hands and arms, 16 denied swelling, and two did not comment. Twenty-two of 50 patients noted being woken up in the middle of the night with headaches; 26 of 50 noted waking up in the morning with headaches. Positional effects were known in 36 patients. Of these, 30 reported having headaches that were worse being reclined, sometimes requiring them to sleep with the head elevated.

**Table 2:**
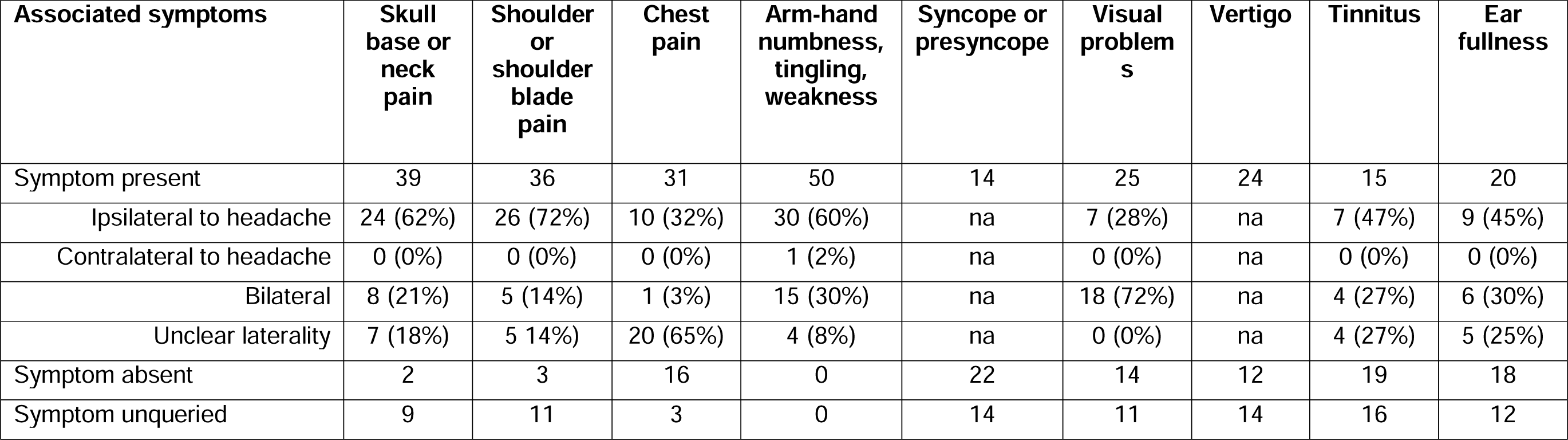
Laterality and associated symptoms with migraine.

| **Associated symptoms** | **Skull**<br>**base or**<br>**neck**<br>**pain** | **Shoulder**<br>**or**<br>**shoulder**<br>**blade**<br>**pain** | **Chest**<br>**pain** | **Arm-hand**<br>**numbness,**<br>**tingling,**<br>**weakness** | **Syncope or**<br>**presyncope** | **Visual**<br>**problem**<br>**s** | **Vertigo** | **Tinnitus** | **Ear**<br>**fullness** |
| --- | --- | --- | --- | --- | --- | --- | --- | --- | --- |
| Symptom present | 39 | 36 | 31 | 50 | 14 | 25 | 24 | 15 | 20 |
| Ipsilateral to headache | 24 (62%) | 26 (72%) | 10 (32%) | 30 (60%) | na | 7 (28%) | na | 7 (47%) | 9 (45%) |
| Contralateral to headache | 0 (0%) | 0 (0%) | 0 (0%) | 1 (2%) | na | 0 (0%) | na | 0 (0%) | 0 (0%) |
| Bilateral | 8 (21%) | 5 (14%) | 1 (3%) | 15 (30%) | na | 18 (72%) | na | 4 (27%) | 6 (30%) |
| Unclear laterality | 7 (18%) | 5 14%) | 20 (65%) | 4 (8%) | na | 0 (0%) | na | 4 (27%) | 5 (25%) |
| Symptom absent | 2 | 3 | 16 | 0 | 22 | 14 | 12 | 19 | 18 |
| Symptom unqueried | 9 | 11 | 3 | 0 | 14 | 11 | 14 | 16 | 12 |

All 50 patients underwent chest CT, in which there were 26 cases of narrowed subclavian or innominate veins equally split between the right and left sides (**Table 3**). Other findings relevant to the diagnosis included three cases of cervical ribs, three cases of thymic hyperplasia causing mediastinal crowding, a thoracic schwannoma impinging on the subclavian vein of the symptomatic arm, and one aberrant 2^nd^ rib insertion into the sternum on the symptomatic arm. MRI of the brain was available in 36 patients and head CT in 26 patients, some of whom had both. There were four cases of cerebellar ectopia, two strokes that occurred after the onset of the migraines, and one case of demyelinating disease. Fifteen patients underwent digital subtraction venography. All confirmed severe large vessel venous stenosis (≥50%) in the subclavian vein. Thirteen of these patients also had the internal jugular vein interrogated; 12 had concurrent internal jugular vein stenosis of ≥50%; 6 were stenosed ≥75%. Seven underwent additional arteriography with two showing mild concurrent subclavian artery stenosis (≤25%).

**Table 3:** Imaging characteristics of chronic migraine TOS patients.

| **MRI brain with or without contrast (n=36)** |  | **CT chest with contrast (n=50)** |  |
| --- | --- | --- | --- |
| Normal | 24 | Right SCV or BCV stenosis | 13 |
| Small vessel disease | 4 | Left SCV or BCV stenosis | 13 |
| Cerebellar ectopia | 4 |  |  |
| Occipital lobe stroke | 1 | Normal | 13 |
| Cerebellar stroke | 1 | Non-diagnostic (poor contrast) | 3 |
| Middle cerebral artery aneurysm | 1 |  |  |
| Demyelination | 1 | *Structural abnormalities* |  |
| Cervical spine syrinx | 1 | Cervical rib | 3 |
|  |  | Thymic hyperplasia | 3 |
| **CT head without contrast (n=26)** |  | Thoracic schwannoma | 1 |
| Normal | 21 | Anomalous rib insertion | 1 |
| Arachnoid cyst | 1 |  |  |
| Parietal cyst | 1 | *\*SCV=subclavian vein* |  |
| Middle cerebral artery aneurysm coiled | 1 | *\*BCV=brachiocephalic vein* |  |
| Vertebral occlusion | 1 |  |  |
| Cerebellar ectopia | 1 |  |  |
| No MRI or CT head | 5 |  |  |

Of the 50 patients, 15 had a lumbar puncture with CSF opening pressure measurements. Pressures ranged from 14-36 cmH20, mean 21.8+/-6.9 cmH20, median 21.0 cmH20. Only one patient in the series exhibited papilledema. This patient developed migraine headaches and TOS after a T1 schwannoma resection. She had a pressure of 36 cmH20.

Patients had tried a mean of 4.0+/-2.4 (median number tried = 4, range 0-11) preventative migraine medical treatments (antihypertensives, anticonvulsants, antidepressants, Botox, CGRP inhibitors) before starting treatment for TOS. Patients who eventually had surgery showed a trend towards having tried more medical treatments for migraines than those who did not have surgery (4.5+/-1.9 vs 3.1+/-2.4 medications, two-tailed *p-value* 0.0885).

All patients had a course of physical therapy with a total of 32 patients eventually undergoing surgical decompressive procedures during the observation period. The same patient could undergo more than one procedure. Twenty patients underwent bilateral 1^st^ rib resections with scalenectomy, five right-sided 1^st^ rib resections with scalenectomy, six left-sided 1^st^ rib resections with scalenectomy, five additional separate scalenectomies, four pectoralis minor tenotomies, two vein patches (one internal jugular vein, one subclavian vein), and 13 a trial of PTV. All TOS surgical treatments were done to improve arm symptoms rather than for headaches despite improvement in headaches being of primary interest in this study. Patients were followed for an average of 7.2+/-5.2 months (median 7 months, range 1-19 months) after their last procedure. The follow-up was done by either a vascular surgeon or a neurologist. One patient in this series developed subclavian vein thrombosis that led to a pulmonary embolus before she had her 1^st^ rib + scalenectomy surgery. Two patients had posterior circulation strokes with cryptogenic sources before being diagnosed with TOS.

Risk factors for the development of TOS are detailed in **(Supplemental Table 1)**. Thirty-seven patients had a history of trauma to the head, neck, or chest that ranged from overuse injury to the instrumentation of the neck or chest. Some patients had more than one risk factor.

### Treatment response

Twenty-eight of 41 patients in whom post-treatment clinical status was known reported ≥50% better headache control after treating their TOS. Twelve of these 28 patients reported 100% headache relief. Overall headache relief in the 41 patients averaged 72+/-26.7% (range 50-100%) by their global assessments. Four reported no improvement. Two of these four patients had no improvement in headache but had resolved or majority improvement in syncopal spells. One patient had no improvement in headache but had resolved supine chest pain and palpitations.

Patients whose symptoms did not improve were typically in the middle of treatment at the time data were summarized and were waiting for surgery on the contralateral side. Nine patients did not follow up after treatment was initiated to know their response. The two patients with hemiplegic migraines had hemiplegic symptoms resolve with surgery (cervical rib removal in one, bilateral 1^st^ rib resections in one). In seven patients, headaches recurred after a period of 1-5 months on the contralateral side which prompted surgery of the contralateral side.

### Survey participants

Of the 50 patients invited to take the survey, 26 started the survey. Of these, 24 completed enough sections to allow data analysis. Of the remaining 24 who did not take the survey, one patient declined to participate, one had an inactive email address, and 22 did not respond. To determine whether there were any systematic biases in participants (n=26) and non-participants (n=24), the general characteristics of each group were compared (**Supplemental Table 2)**. Demographic features were similar including sex distribution, age, years of migraine, and years of chronic migraine, but were different in duration of care (1.5+/-0.9 years participants vs 1.0+/-0.9 years non-participants, *p=0.0276*) and a greater proportion of participants who had started the survey who had undergone surgery (19 of 24 participants =79%) than non-participants (11 of 24 =49%).

### Clinical features of survey participants

Eighteen of 24 patients who completed the survey reported baseline chronic daily headaches with no headache-free days of which 11 patients had headaches that were chronic from the onset. The most frequent area of maximal pain was the skull base, with the most frequent description of the pain being, “pressure, head exploding,” followed by “throbbing.” The relationship between the onset of headache and limb symptoms of TOS was answered by 23 participants. Limb symptoms with headaches occurred either “always,” or “sometimes,” with headaches in 87% of participants **Table 4**.

**Table 4:** Features of headache quality and temporal qualities in survey responders.

| **Headache features** | ***Participants***<br>***(n=24)*** | **%** |
| --- | --- | --- |
| **Laterality** |  |  |
| Predominantly unilateral | 15 | 63 |
| *Right* | 10 | 42 |
| *Left* | 5 | 21 |
| Alternating unilateral | 4 | 17 |
| Always bilateral | 5 | 21 |
| Unsure | 1 | 4 |
| **Area of maximum pain** |  |  |
| Skull base | 11 | 46 |
| Forehead | 5 | 21 |
| Vertex | 4 | 17 |
| Temples | 4 | 17 |
| Around ear | 1 | 4 |
| **Headache description** |  |  |
| "Pressure, head exploding" | 10 | 42 |
| "Throbbing" | 5 | 21 |
| "Pressure, head squeezing" | 5 | 21 |
| "Sharp shooting" | 3 | 12 |
| "Dull" | 2 | 8 |
| **Years between onset of migraine and arm symptoms** | ***Participants***<br>***(n=23)*** | **%** |
| within 1 year | 10 | 43 |
| 1-5 years | 6 | 26 |
| 5-10 years | 1 | 4 |
| >10 years | 6 | 26 |
| **Relative onset of migraine to arm symptoms in years** |  |  |
| Migraine onset before arm symptoms | 17 | 74 |
| Arm symptoms onset before migraine | 5 | 22 |
| Migraine and arm symptoms onset same time | 1 | 4 |

| **Synchronicity of headache and arm symptoms for each migraine episode** |  |  |
| --- | --- | --- |
| Always together | 4 | 17 |
| Sometimes together | 16 | 70 |
| Never together | 3 | 13 |

### Treatment responses from survey participants

Participants rated the change in their headache burden from before to after treatment for TOS with reports corroborated with the clinical record. The average number of headache days per month decreased from 18.3+/-8.6 to 11.1+/-10.8 (p<0.0016) with the average severity of headache on a 0-10 scale decreasing from 7.8+/2.5 to 5.4+/-2.9 (p<0.00079). The average number of visits to an urgent care or emergency room per year decreased from 3.6+/-4.0 to 0.71+/-1.3, p<0.0029). For the 19 of 24 survey participants with completed data and who had undergone any surgical procedures, the survey was reported at a mean of 10.0+/-6.1 months (median 9 months, range 0.5-20.7 months) after their last surgical procedure. Of note, patients did not have to have completed all planned surgical procedures to participate in the survey.

Eighteen patients made responses on a 5-point Likert scale of “strongly disagree (-2),” to “strongly agree (+2),” for general statements assessing their well-being and physical capabilities at home and at work, both before and after their TOS diagnosis and intervention. The mean change for “I am fully functional at work,” was positive (mean 1.22+/-1.48; range 0-4). Similarly, “I am fully functional at home,” indicated a positive change (mean 1.17+/-1.34; range 0-3) (**Figure 3**).

**Figure 3:**
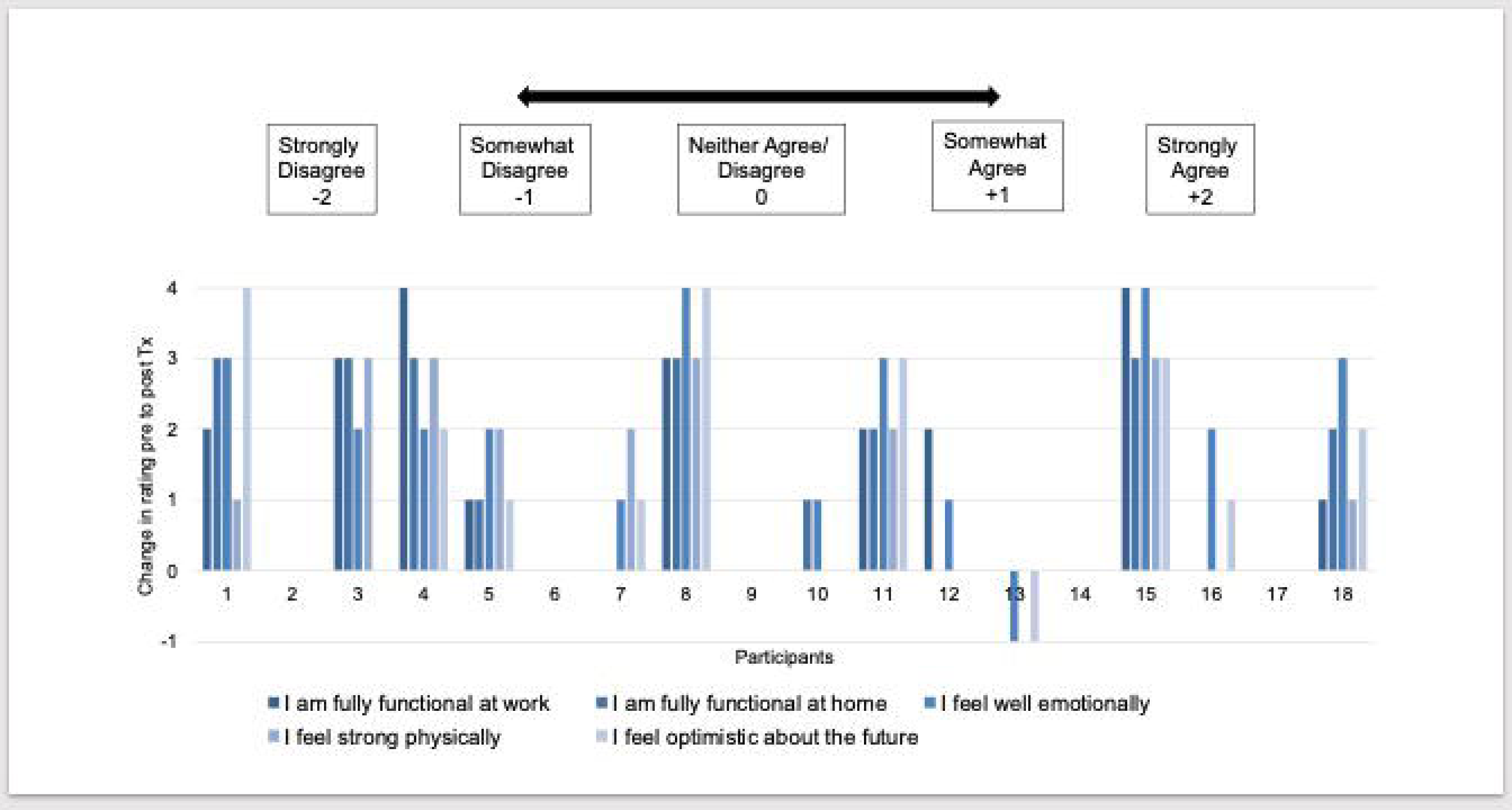
Change in patient participants’ rating of statements related to their functional capabilities before and after treating underlying TOS. Scales go from Strongly Disagree (-2) to Strongly Agree (+2) for a maximum possible change of 4 points.

## DISCUSSION

This study reports the real-life clinical experience of how treating concurrent TOS can alleviate chronic migraine headaches in patients with these dual diagnoses. It more than doubles the number of patients reported in the literature for which this association is important and could thus buttress the rationale for future controlled trials for this underappreciated association.

Thoracic outlet syndrome as a contributor to chronic migraines should be considered in patients with certain clinical features. Our study indicates that these features include: 1. Headaches ipsilateral to myofascial symptoms including skull base, neck, and shoulder pain, along with arm and hand symptoms of numbness, tingling, and weakness 2. Headaches associated with edema of the face, arm, and/or hand, 3. Symptoms worse in recumbency or that frequently wake a person from sleep, 4. Headaches developing in the context of neck and chest trauma, including postural strain, 5. Headaches that are chronic within one year of onset, including NDPH. While no feature is specific to TOS, the combination of supporting features should raise consideration for an underlying structural disorder such as TOS contributing to migraine pathology.

In this study, all patients met CORE TOS SVS criteria for both nTOS and vTOS through imaging and clinical exams. The nTOS component served as the signal that the thoracic outlet was compromised whereas the vTOS component explained the clinical features related to the migraines. It is possible that vTOS alone could contribute to migraines without leading to arm problems, but these patients would be difficult to find unless routine screening for TOS was performed in all migraine patients. It remains for a future study to determine whether this is reasonable.

The anatomy of the venous system of the head, neck, and thoracic outlet is consistent with most clinical features. Subclavian vein obstruction in vTOS impedes drainage from the external jugular vein and potentially the internal jugular vein when the obstruction is near the confluence of the innominate vein. During digital subtraction venography, the external jugular vein was frequently noted to be overdeveloped with exuberant collaterals (**Figure 2B**). This may lead to elevated venous pressure in tributaries upstream from the compression that can lead to neck pain and skull-base pressure that triggers a migraine ^21, 22^. The areas drained by these veins include the skull base, lateral neck, periauricular, and suprascapular areas--all areas in which both swelling and pain have been noted in these patients. Because emissary veins carrying blood from the internal jugular veins collateralize with the external jugular vein at the skull base, subclavian vein obstruction may potentially impact flow in the internal jugular vein leading to downstream effects on drainage from the cavernous sinus around the eye and the superior and inferior petrosal sinuses that drain the inner ear ^23, 24^. The superior petrosal sinus sits over the trigeminal nerve and sometimes wraps around it creating a potential mechanism for the direct triggering of facial pain with venous congestion ^25^. Moreover, the increasingly recognized role of the jugular venous system in regulating intracranial pressure may be a mechanism for headaches, particularly those triggered by postural changes ^26–28^. Notably, most patients experienced worse headaches in the supine position when venous outflow is most dependent on the jugular pathway as opposed to the paravertebral venous plexus that carries greater venous outflow in the upright position ^26–28^. Almost half of the patients in this series noted being woken up in the middle of the night by headaches. Further, the re-routing of venous blood through intrathoracic veins in upper extremity venous compressive disorders could be an important factor in the associated symptoms of chest pain and palpitations, particularly those worsened by supine positioning when venous pressure is high^9, 10, 29, 30^.

### Special cases

The two patients with hemiplegic migraines are of particular interest. One mechanism of hemiplegic symptoms in vTOS relates to the parallel path of venous outflow between the paravertebral venous plexus and the jugular system which bidirectionally shunts with postural changes ^28, 30–32^. Compression of the jugular system shunts venous blood to the vertebral veins which drain the cervical spinal cord ^33^. Over-shunting of venous blood due to either fixed or frequent obstruction of the jugular venous system could lead to venous congestion and reduced perfusion pressure of the ipsilateral spinal cord. This can trigger hemiplegic symptoms. In the patient with the cervical rib, TOS decompression also likely removed a component of subclavian artery compression causing limb ischemia.

Two patients had a history of cryptogenic embolic strokes. While one patient had a history of hyperlipidemia, she was only in her 30s and otherwise in excellent health when she developed a caudal cerebellar hemispheric stroke. The other patient was in her 40s and developed an occipital stroke in the setting of a particularly bad migraine headache. Strokes have been reported in TOS, generally in the context of obstructed flow through a stenosed subclavian artery and stagnation of blood in an adjacent aneurysm ^34, 35^. Neither of these situations applied to our patients but we note that one of these patients had complete resolution of her headaches after addressing her nTOS and vTOS.

### Limitations

This report expands the scope of clinical features reported on patients with both chronic migraine and TOS. It is not a controlled clinical trial, however, and thus has limitations. Baseline clinical features were reported from chart review over a limited time period with different durations of follow-up. There was no natural control arm for this study other than showing that standard migraine strategies had failed in these patients. Compared to non-participants, there were proportionately more participants who had responded to the questionnaires who had undergone surgical procedures, who came to follow-up appointments, and who had reported having better headache control at the last clinical follow-up. Thus, the true percent improvement may be lower than if all patients had responded. Mitigating these biases is that there were no significantly different baseline characteristics in clinical features between responders and non-responders to the questionnaire. Further, there was a counter bias towards poor response in that some patients were in the middle of treatment during their reporting period rather than at the end of treatment.

Ideally, a longitudinal approach could be taken to establish recurrent surveys over the course of several years with the caveat that more demanding studies may lead to reduced participation and still select for patients with better responses. Finally, the sample size is small relative to other headache and vascular studies but a report of 50 patients with combined TOS and chronic migraine represents the largest case series presented on this association to date in the world English literature. With the recognition that these disorders are more related than currently suspected, these cases may appreciably increase with time. This case series may help identify patients with TOS and migraine for future trials.

## CONCLUSION

Chronic migraines can be important manifestations of TOS that can improve with the treatment of TOS. TOS should be suspected when neck pain, shoulder pain, and limb paresthesia or swelling are present, particularly when ipsilateral to the headache, and when headaches are described as a pressure that is worse with recumbency. We propose that this association is much more common than is usually appreciated due to the frequency of head, neck, and chest trauma from overuse injury including some iatrogenic causes such as central lines or neck surgery. TOS- associated headaches may be chronic from the onset and should be considered in the differential in patients who meet the criteria for migraine and NDPH. Identifying secondary causes of headaches such as TOS is of critical importance to avoid losing a window of opportunity to treat before fibrosis and venous stenosis become permanent and drastic measures such as vein patching are needed. The consideration of TOS as a possible contributor to chronic migraines may help address at least one segment of the chronic migraine population and expedite appropriate therapy. We hope that this report will help justify research resources to study the association between TOS and migraine more comprehensively.

## Ethics approval and consent to participate

Institutional approval was obtained from the Saint Francis Health Systems Institutional Research Ethics Board (Tulsa, OK), protocol number 2250-19. A chart review of all patients seen by the principal investigator over the course of 3.5 years was reviewed for a diagnosis of thoracic outlet syndrome. Data are presented anonymously. Patients in the questionnaire study provided information with participant IDs.

They were allowed to withdraw from the study at any time. All data were treated as confidential.

## Consent for publication

Data from chart review are presented anonymously without personally identifying information. There are no identifying features in the images. Patients who were in the questionnaire study provided consent for participation and publication.

## Availability of data and materials

The dataset supporting the conclusions of this article is available in the Harvard Dataverse: https://dataverse.harvard.edu/dataset.xhtml?persistentId=doi:10.7910/DVN/Y4AEPL

## Authors’ contributions

LR performed data analysis, drafted the paper, and reviewed the interim and final manuscripts. JW provided patient data and reviewed the interim and the final manuscript. SA provided patient data and reviewed the interim and the final manuscript. YHC conceived the study, provided patient data, administered the questionnaire study, and reviewed all drafts of the manuscript.

## Data Availability

Availability of data and materials: The dataset supporting the conclusions of this article is available in the Harvard Dataverse.

https://dataverse.harvard.edu/dataset.xhtml?persistentId=doi:10.7910/DVN/Y4AEPL

## Acknowledgments

The authors thank the patients who participated in the study. The authors received no compensation outside of routine clinical care for these patients. There was no external funding for this study and no involvement of 3^rd^ party institutions in the collection of data or development of this manuscript.

