## Supplemental Table 1 for "Treatment of thoracic outlet syndrome to alleviate chronic migraine headache"

**Supplemental Table 1: Risk factors for TOS**

| **Risk factors for TOS** |  |
| --- | --- |
| Known overuse mechanism | 7 |
| Blunt trauma to head/neck/chest | 6 |
| Breast implants | 5 |
| Cervical spine fusion | 4 |
| Thymic mass | 3 |
| Cervical ribs | 3 |
| Clavicle fracture | 2 |
| History of central lines in neck | 2 |
| Hypermobility disorder | 2 |
| Immediate post-op | 2 |
| Anomalous rib insertion | 1 |
| Thoracic schwannoma | 1 |
| Unknown | 18 |
