## Supplemental Table 2 for "Treatment of thoracic outlet syndrome to alleviate chronic migraine headache"

**Supplemental Table 2: Differences in survey participants and non-participants**

|  |  | **Participants (n=26)** |  | **Non-participants**  **(n=24)** |  | p-value |
| --- | --- | --- | --- | --- | --- | --- |
|  |  | *Mean +/- s.d.* | *Median (range)* | *Mean +/- s.d.* | *Median (range)* |  |
| Age at presentation | | 43.2+/-13.3 | 44 (17-66) | 44.8+/-12.3 | 44 (19-78) | *0.662* |
| Age onset migraine | | 23.4+/-14.9 | 22 (5-62) | 24.3+/-15.2 | 25 (5-48) | *0.832* |
| Age onset chronic migraine | | 34.9+/-14.3 | 35 (5-62) | 34.6+/-12.6 | 37.5 (5-59) | *0.946* |
| Duration chronic migraine | | 8.4+/-9.5 | 6.5 (0.33-41) | 10.2+/-15.5 | 5.3 (0.3-73) | *0.620* |
| Duration follow-up | | 1.5+/-0.9 | 1.5 (0-3.2) | 1.0+/-0.9 | 0.9 (0-3.1) | *0.0276* |
